# Enhancing Leptospirosis Diagnosis through an integration of Machine Learning with Classification of Microscopic Agglutination Images

**DOI:** 10.1101/2024.10.07.24315068

**Authors:** Norhasmira Mohammad, Murnihayati Hassan, Siti Nur Zawani Rosli, Natasya Amirah Tahir, Nurul Azmawati Mohamed, Khairunnisa Mohd Sukri, Liyana Azmi

## Abstract

Leptospirosis, a widespread zoonotic disease, poses substantial challenges to global public health. In Malaysia, leptospirosis is an endemic disease, with annual cases peaking during the monsoon season. The microscopic agglutination test (MAT) is the gold-standard serological method for confirmation of leptospirosis. However, it is labour-intensive and time-consuming, as it relies on the subjective interpretation of medical lab technicians. This study investigates and describes the development of a semi-automated workflow for leptospira screening by integrating a TensorFlow and custom-designed Keras-based Deep Convolutional Neural Network (DCNN) with conventional MAT. We used a dataset of 442 positive and 442 negative MAT images, which consisted of a mixture of leptospira serovars to train the model. We then subjected our model to hyperparameter tuning, where we adjusted various settings to optimise the model’s performance. These settings included the number of convolutional layers, filters, kernel sizes, units in dense layers, activation functions, and the learning rate. We then tailored several convolutional layers to find the optimal balance between model complexity and performance. Verification of our tested model compared to the control samples (verified patient MAT results) achieved the following metrics: a Precision score of 0.8125, a Recall of 0.9286, and an F1-Score of 0.8667. Combining our model with the current Malaysia leptospira workflow can significantly speed up, reduce inaccuracies and improve the management of leptospirosis. Furthermore, the application of this model is practical and adaptable, making it suitable for other labs that observe MAT as their leptospira diagnosis. To our knowledge, this approach is Malaysia’s first hybrid diagnostic approach for leptospira diagnosis. Scaling up the dataset would enhance the model’s accuracy, making it adaptable in other regions where leptospirosis is endemic.

## Introduction

Leptospirosis, a zoonotic disease, is transmitted from the urine of host animals to humans through skin breaks or mucous membranes. This potentially severe ailment is particularly prevalent in developing countries such as Malaysia, India, Sri Lanka, and Brazil where thousands of cases are reported annually (1). The disease exhibits a spectrum of symptoms ranging from mild to severe, often leading to multiorgan failure and, in extreme cases, death. Given the non-specific clinical manifestations of leptospirosis, which can resemble other infections such as influenza and dengue, a combination of good clinical judgment and reliable diagnostic tools becomes crucial. In Malaysia, leptospirosis is an endemic and notifiable disease since 2010 and has been recording an increased incidence rate, rising from 1.03 cases per 100,000 to 30.2 cases per 100,000 (2). Alarmingly, the disease has also resulted in a notable mortality rate of 0.45 per 100,000 persons (3). Urgent attention and proactive measures which includes robust diagnostic methods to promptly identify and manage cases, are necessary to prevent spread and potential fatalities. However, diagnosing leptospirosis presents several challenges, leading to gaps in disease management.

Early detection during acute stage is dependent upon on rapid diagnostic test (RDT). However, RDT tests are prone to false negative (FN) and false positive (FP) results, as it relies on the antigen selection for the design of the kit. RDT specificities are compromised due to cross-reactivity with other antibody of infectious origin. In addition, timing of the specimen is very crucial for serological-based RDT especially those relying on IgM alone. Utility of *Leptospira* PCR for early laboratory confirmation is highly recommended but the test is not readily accessible in endemic region with resource limitation (4). Therefore, gold standard serological method based on leptospirosis microscopic agglutination test (MAT) remain as the best confirmatory test of choice despite of delay in positivity during the stage of the illness. MAT surpasses specificity of other serological based testing method as the patient serum will be directly reacted with a panel of live *Leptospira* serovars.

Other methods for leptospirosis diagnosis include antibody detection such as Enzyme-Linked Immunosorbent Assay (ELISA), which is commonly employed for initial screening due to their ease of use and availability (5). One major concern with serological tests is the issue of cross-reactivity. Antibodies produced during other infections, especially those caused by closely related bacteria, can show cross-reactivity with false antigens (6). This cross-reactivity can lead to FP results, complicating the diagnosis and potentially causing mismanagement of cases. Moreover, the timing of sample collection is critical for serological testing. The optimal time to collect samples is during the acute phase of the disease, typically within the first ten days of symptom onset (7).

One significant challenge lies in the microscopic characteristics of leptospires, as they cannot be observed under an ordinary light microscope direct from clinical specimens (4). Instead, a specialised darkfield microscopy is required. Furthermore, direct examination of blood and urine using darkfield microscopy has limitations in both sensitivity and specificity (8), making it less reliable for definitive diagnosis. In the other hand, MAT utilises different types of serovars reacted with patient’s serum at serial dilution to observe for presence of total neutralizing antibody. MAT allows identification of potential infecting serovar thus, contributing to epidemiological surveillance in human leptospirosis. However, the interpretation of MAT is complex. Agglutination patterns can vary between samples and serovars thus, require experienced personnel to accurately read and interpret them. In cases with low antibody titers or weak reactions, the interpretation becomes particularly difficult, increasing the risk of FN or FP results.

Incorporating artificial intelligence (AI) to improving leptospirosis diagnosis can be beneficial. One of the most used AI-methods include deep learning (DL) algorithms. DL is a form of machine learning which focuses on training computer models to learn and perform tasks (9). During the ‘training’ period, the algorithm is presented with a large dataset containing inputs and their corresponding correct outputs. The network adjusts its internal parameters, in an iterative process called backpropagation. The backpropagation then comparing the algorithm output to the correct output (validation), calculates the error, and then propagates the error backward to reduce the error. This iterative learning process continues until the network’s performance reaches a satisfactory level.

This research proposes an approach to enhance the diagnostic process for leptospirosis, leveraging advanced technologies such as TensorFlow and the Keras-based Deep Convolutional Neural Network (DCNN). By integrating DL with the conventional MAT, we aim to streamline and expedite the diagnostic process for leptospira diagnosis. The existing diagnostic tests are known for their complexity and prolonged duration, prompting the exploration of a novel strategy utilizing TensorFlow and Keras-based DCNN to enhance the efficiency and speed of leptospirosis diagnosis by accurately assignment of binary outcome of individual MAT screening reaction of individual serovars.

## Methodology

### Dataset

The dataset used in this study comprised 884 images which consists of 442 positive and negative images of leptospirosis from archived images of positive MAT sera from acute febrile illness cases. This collection provides a diverse set for training and evaluating the Deep Convolutional Neural Network (DCNN) implemented with TensorFlow and Keras. The balanced representation of positive and negative reactions of individual serovar reaction ensures the robustness of the model, enabling it to effectively learn and identify key features associated with leptospirosis for enhanced diagnostic accuracy. Each image was generated from acute serum prepared in 1:50 dilution, where a positive MAT reaction is defined by 50% agglutination of the individual serovar in comparison to control (individual serovar without addition of patient’s serum).

### Image pre-processing

All raw JPEG images underwent resizing to ensure uniform dimensions. The raw JPEG images were cropped into 250 * 250-pixel sub-images using ImageJ (10) to contain the same input size and spatial resolution. Secondly, adjustments to brightness and color were made across all images. These cropped binary images were then saved in JPEG format, forming the training dataset that is homogenous dataset for DCNN training.

### Model Development

#### Architecture Selection

The selection of DCNN for this study was driven by its capability to effectively capture intricate features indicative of leptospirosis within the dataset. DCNNs excel in image classification tasks (11), making them particularly suitable for discerning patterns and features present in medical images associated with diseases such as leptospirosis.

#### Hyperparameter Tuning

The configuration of DCNN was fine-tuned using a dataset comprising 442 positive and negative images. This adjustment of the DCNN’s settings, executed through the TensorFlow and Keras frameworks, enhances the model’s proficiency in identifying distinctive features associated with leptospirosis. By leveraging this large and diverse dataset during the hyperparameter tuning process, the DCNN becomes more adept at recognizing subtle patterns and nuances indicative of the presence or absence of leptospirosis.

#### Hyperparameter Optimization

In this study, hyperparameter optimization is performed using the Keras Tuner library and a RandomSearch strategy. Key hyperparameters, including the number of convolutional layers, filters, kernel sizes, units in dense layers, activation functions, and the learning rate, are explored within a defined search space. The Keras Tuner object is configured, and a randomised search is conducted on the training data to identify the optimal hyperparameter set. The optimal parameters are subsequently employed for the construction and training of a new model. This model implementation includes an early stopping callback, which oversees validation loss during training and interrupts the process if improvements level off. The resulting model is evaluated on the test set, presenting metrics such as test loss and accuracy. This automated hyperparameter tuning enhances the model’s generalization and adaptability, ensuring superior performance across diverse datasets.

### Model Evaluation

#### Performance Metrics

The evaluation of the DCNN for identification of positive and negative MAT images involves key performance metrics: accuracy, which measures overall correctness; precision, indicating accuracy in positive predictions; recall (sensitivity), gauging the ability to identify relevant instances; F1-Score, ensuring a balanced assessment; the confusion matrix, offering a visual representation of predictions; the Receiver Operating Characteristic (ROC) curve, illustrating sensitivity vs. specificity trade-offs; and loss, evaluating model prediction errors during training. These metrics collectively provide a comprehensive understanding of the DCNN’s diagnostic performance.

#### Inference on Test Set

The evaluation of the pre-trained DCNN model for leptospirosis MAT diagnostic tool involves testing it on a new set of images. This assessment aims to gauge the model’s effectiveness in predicting positive MAT images based on the 50% neutralisation of free leptospires in comparison to control (Leptospira serovar without addition of patient’s serum). The images undergo preprocessing, including resizing and normalization, before being fed into the model. The model generates predictions, and the resulting binary labels are compared with true labels. Visual representations of the images, accompanied by their predicted classes, enhance the qualitative analysis. Subsequently, computed metrics such as accuracy, F1 score, confusion matrix, and a classification report provide a comprehensive evaluation of the model’s performance on previously unseen data. This process is pivotal in validating the model’s applicability to real-world scenarios and ensuring its reliability in interpretation of MAT images.

## Statistical Analysis

The comparison between the model’s interpretation and the actual MAT results for individual serovar was carried out by contrasting the model’s output with the genuine MAT results performed by medical lab technologists. This comparison was then quantified as a mean error (ME) to determine the direction of the error, whether it was an overestimation or underestimation of the results. The statistical significance of the differences between the same variables from different datasets, two types of statistical tests were employed: the independent t-test and the paired t-test. The level of statistical significance was established at p < 0.05. All statistical analyses were conducted using RStudio software, specifically version 0.97.551 (2009–2012; RStudio, Inc.).

## Results

### Developed Pre-trained Model from Scratch

The best hyperparameter-tuned model is a Sequential Convolutional Neural Network (**S**CNN) designed to process input images resized to 128×128 pixels. Originally, the images were 250×250 pixels, but the reduction in size to 128×128 was intentionally done to expedite computational time while maintaining effective diagnostic capabilities for leptospirosis. The architecture consists of four convolutional layers (Conv2D) with max-pooling layers (MaxPooling2D) in between, employing multiple filters of various sizes. Dense layers are included for classification, resulting in a total of 2,164,193 trainable parameters. This optimised configuration enhances the model’s ability to recognise relevant features in leptospirosis diagnosis, providing an efficient balance between computational efficiency and diagnostic accuracy. **Table 1** illustrates the architecture of the DCNN developed through the algorithm’s hyperparameter tuning process.

**Table 1:** The DCNN architecture generated by the algorithm after hyperparameter tuning.

| Model: “sequential” |  |  |
| --- | --- | --- |
| Layer (type) | Output shape | Parameters |
| Conv2D_1 | (None, 124, 123, 144) | 109,44 |
| MaxPooling2D_1 | (None, 62, 62, 144) | 0 |
| Conv2D_2 | (None, 56, 56, 112) | 790,384 |
| MaxPooling2D_2 | (None, 28, 28, 112) | 0 |
| Conv2D_3 | (None, 24, 24, 144) | 403,344 |
| MaxPooling2D_3 | (None, 12, 12, 144) | 0 |
| Conv2D_4 | (None, 8, 8, 80) | 288,080 |
| MaxPooling2D_4 | (None, 4, 4, 80) | 0 |
| <sup>a</sup>Flatten (Flatten) | (None, 1280) | 0 |
| <sup>b</sup>Dense_1 (Dense) | (None, 432) | 553,392 |
| <sup>b</sup>Dense_2 | (None, 272) | 117,776 |
| <sup>b</sup>Dense_3 | (None, 1) | 273 |
| Total parameters: 2,164,193 |  |  |
| Trainable parameters: 2,164,193 |  |  |
| Non-trainable parameters: 0 |  |  |

<sup>a</sup>Transformation of a two-dimensional (2D) output into a one-dimensional (1D) output.
<sup>b</sup>Hyperparameter used to establish connections between every neuron in the current layer and every neuron in the previous layer to compute the output.

### Performance evaluation of the DCNN model

The classification results for leptospirosis, based on the developed model, exhibit promising performance. **Table 2** shows the accuracy from each evaluation metrics. Precision, which measures the accuracy of positive predictions, was observed to be 0.8125, indicating that approximately 81.25% of instances predicted as positive were accurate.

**Table 2:** Performance metrics.

| Precision | Recall | F1-Score |
| --- | --- | --- |
| 0.81 | 0.93 | 0.87 |

The Recall metric, quantifying the model’s effectiveness in capturing actual positive instances, stands at 0.9286 (92.86%), which suggests a good sensitivity matric for identification of leptospirosis cases. In medical diagnostics, achieving a high Recall is crucial to minimise the risk of false negatives and ensure comprehensive coverage of positive cases. The F1-Score, which combines both Precision and Recall into a single metric, is calculated to be 0.8667. This balanced measure signifies a harmonious trade-off between Precision and Recall. A higher F1-Score is indicative of a model that excels in both precision and sensitivity, making it well-suited for the specific context of leptospirosis classification.

Our confusion matrix (**Fig. 1**) showed a breakdown of the model’s predictions. The top-left element (Fig 1A) represents 75 images which was correctly identified as true negative (TN) for leptospirosis, while the bottom-right element (Fig 1B) represents 78 true positive (TP) leptospira samples. Notably, there were instances of misclassifications, as indicated by the top-right (Fig 1C), which correspond to 18 samples identified as FN and bottom-left (Fig 1D), which identified 6 samples as FP. Based on the balanced accuracy, the model shows an equal measure of precision and sensitivity.

**Fig. 1.**
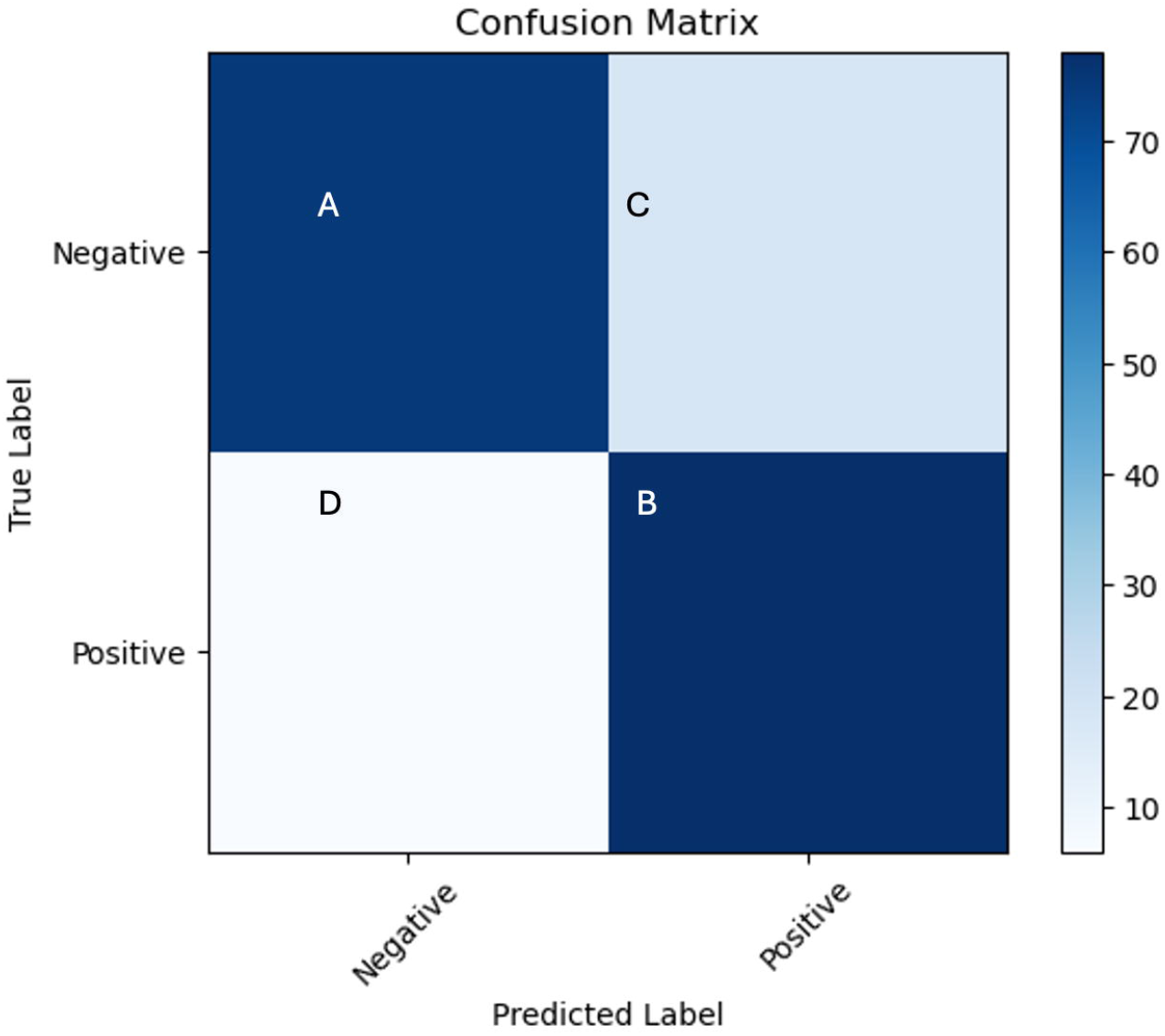
Confusion matrix. (A) TN samples with 75 images, correctly sorted. (B) TP samples with 78 images, correctly sorted. (C) 18 FN samples and (D) 6 FP samples.

We measured the model’s ability to discriminate between positive and negative instances by plotting the Receiver Operating Characteristic (ROC) curve, which is a graphical representation of a binary classification model’s performance across different decision thresholds. Our AUC-ROC value of 0.92, (Fig 2) demonstrated a strong discriminatory power and a high probability that the model will correctly rank a randomly chosen positive instance higher than a randomly chosen negative instance.

**Fig. 2.**
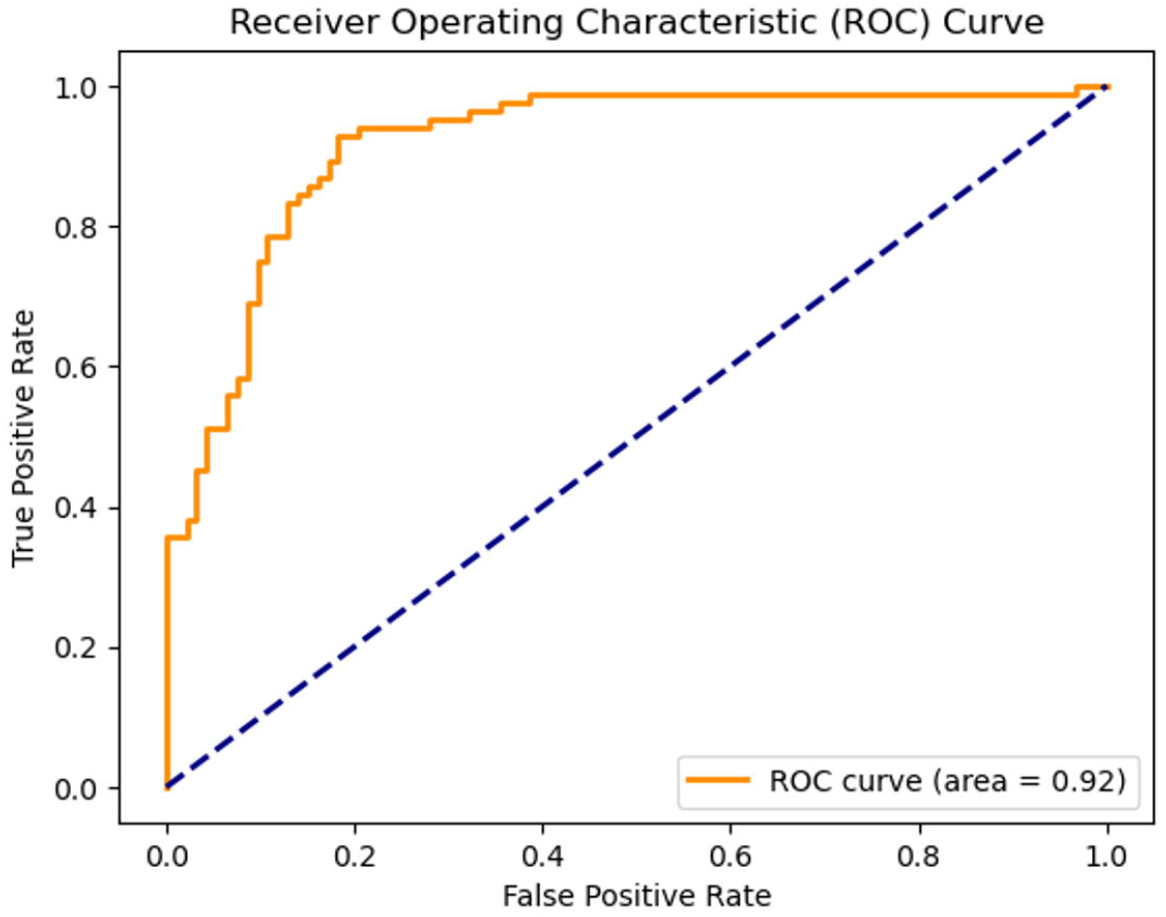
AUC-ROC. (A) TN samples with 75 images, correctly sorted. (B) TP samples with 78 images, correctly sorted. (C) 19 FN samples and (D) 6 FP samples.

### Model Predictions

The inference results (**Table 3**), underscores the performance of the model on the test set. Achieving a perfect accuracy score of 1.0 indicates that the model accurately predicted all instances within the test set. Additionally, the F1 score, a comprehensive metric combining precision and recall, reached its maximum value of 1.0, reflecting an optimal balance between the two and highlighting a model with both high precision and high recall.

**Table 3:** Classification report. The precision, recall and F1-score of the model was tested for its accuracy, macro, and weighted averages.

| Class | Precision | Recall | F1-score |
| --- | --- | --- | --- |
| 0 | 1.0 | 1.0 | 1.0 |
| 1 | 1.0 | 1.0 | 1.0 |
| **Overall assessment** |  |  |  |
| **Accuracy** |  |  | 1.0 |
| **Macro average** | 1.0 | 1.0 | 1.0 |
| **Weighted average** | 1.0 | 1.0 | 1.0 |

Our confusion matrix revealed considerably good performance, with no misclassifications observed. Each element on the diagonal of the matrix represents correct predictions, demonstrating that all instances were accurately classified into either class 0 or class 1. The classification report demonstrates the model’s performance. Precision, recall, and F1-score values for both classes (0 and 1) are all 1.0, which demonstrates a reasonable performance for each class. The macro and weighted averages, considers the average performance metric and contribution of each class based on its proportion in the dataset, respectively. Both macro and weighted averages indicated scores of 1.0.

Image prediction and class sorting, the input images are sorted based on pixel values to classified into positive and negative images in concordance with manual interpretation of MAT reaction. If an image had a low count of pixel values, the image would be predicted and labeled as class 0, which is denoted as class negative and vice versa. Based on the outcome prediction classes (Fig 3), our model showed reasonable sorting capabilities and presents itself as a tool that can assist technologists in making preliminary diagnosis for leptospirosis.

**Fig. 3.**
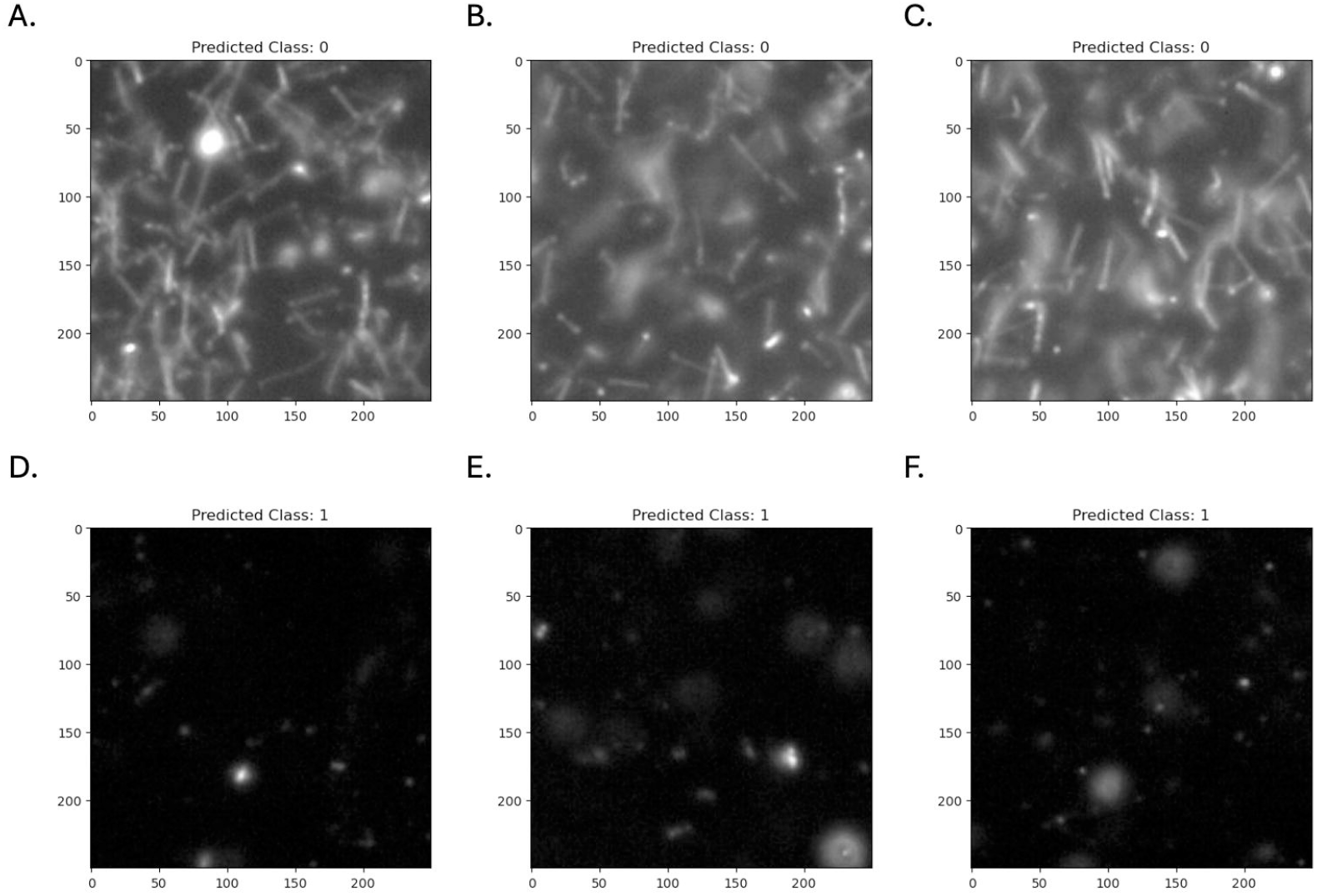
Images with prediction classes post model inference. (A-C) Positive samples, denoted as predicted class 0 and (D-F) negative samples, denoted as predicted class 1 3 Image classification after model inference.

## Discussion

The Microscopic Agglutination Test (MAT) involves evaluating total antibodies present in the blood of suspected patients as they interact with live leptospiral antigens. This procedure entails incubation of 7 to 12-day-old leptospiral serovars to serially diluted patient serum antibodies from patients prepared in a microtiter plate format (12). The interaction between antigens and antibodies is quantified by observing the highest serum dilution causing agglutination under a dark-field microscope, resulting in a 50% reduction of live Leptospira spp compared to the control serovar without the addition of the patient’s serum (13). The traditional manual interpretation of MAT results is prone to human errors due to several factors. Firstly, the subjective nature of visually assessing agglutination patterns can lead to inconsistencies between different interpreters (14). Additionally, human factors such as distractions, fatigue, and other influences can negatively impact the accuracy of manual interpretation. Furthermore, the complexity of the test, involving a variety of serovars, increases the cognitive load and risk of errors, thereby requiring trained expertise (8).

These challenges limit the scalability of the test, especially when processing a large number of patient samples. Despite these limitations, the MAT remains the gold standard for leptospirosis diagnosis, particularly in endemic regions like Malaysia (15). Accurate interpretation necessitates that medical laboratory technologists possess the necessary technical skills, making MAT a method exclusive to specialised infectious disease centers (16), despite its widespread use in endemic regions (17)

There are other alternatives to MAT for leptospirosis diagnosis, such as the ELISA. However, these methods do not address the specificity need of identifying infections based on the serovar of the leptospire (17). Thus, to address the gap in time consumption and improve diagnostics for leptospirosis, we have developed a semi-automated workflow using a customised DCNN model that was tailored to assist leptospirosis diagnosis using MAT images. This approach not only expedites the diagnostic process but also ensures consistency, offering a valuable tool for healthcare professionals in managing leptospirosis cases effectively (18).

Several teams have devised automated methods for leptospira diagnosis (19). In this regard, our method did not adopt a similar machine learning framework nor used a pre-trained model; however, we trained the DCNN model from scratch and constructed new layers and hyperparameter tuning to suit our dataset and output requirements.

The classification results highlight the model’s robust performance, demonstrating a precision of 0.81 in accurately identifying most positive cases. Additionally, a recall score of 0.93 indicates the model’s effectiveness in capturing most leptospirosis cases, minimising FN. The F1-Score, a balanced metric at 0.87, illustrates the model’s efficiency in both precision and recall, crucial for reliable disease prediction.

During inference with the test dataset, a perfect accuracy of 100% was achieved, suggesting potential overfitting. To address this concern, we first investigated instances where the model incorrectly predicted positive cases. Identifying common characteristics or patterns in these FP can guide adjustments to the model architecture, feature extraction, or data preprocessing. By analysing these misclassifications, we can better understand the limitations of the model and implement targeted improvements to enhance its generalisability. Notably, for our model setup, we used an 80:20 ratio of the dataset for training and validation, respectively. Increasing the number of images at the start of the project could enhance the size of the validation set, potentially mitigating overfitting. Therefore, a larger sample size might help reduce overfitting observed in the validation stage (20).

Moreover, depending on clinical implications, adjusting the decision threshold of the model to focus on better sensitivity or specificity is crucial. This adjustment can be tailored to align with desired clinical outcomes and the consequences of FP or FN (21). Achieving an optimal threshold requires fine-tuning TP, TN, FP and FN metrics. By adjusting the threshold, we essentially control how the model classifies its predictions, which directly impacts these metrics. For our model, since our validation showed 100% accuracy, we did not modify the sensitivity and specificity thresholds. In the future, when larger datasets are utilised for model training and validation, the accuracy may no longer be 100%. In this event, we would modify the sensitivity and specificity thresholds to optimise the balance between TP and TN scores, thereby managing the rates of FN and FP.

Finally, validating the model on diverse datasets is crucial to ensuring its generalisation across different contexts (22). On this avenue, considering our variable serovars of leptospira used, we believe that our dataset is satisfactory and allows generalisation for a general screening of leptospirosis. This comprehensive evaluation will provide insights into the model’s robustness and aid in optimising its performance for real-world applications.

In addition, the 100% accuracy observed during inference prompts intriguing considerations. While high accuracy is desirable, careful examination is necessary to ensure robust generalisation. Factors contributing to this may include the inference dataset closely aligning with the training data in distribution, potential over-fitting capturing noise or specific patterns, and a small or less diverse inference dataset leading to memorisation of specific examples rather than learning broader patterns. Ensuring the quality and consistency of the inference dataset is vital, with high-quality images closely aligning with trained features for more accurate predictions. To address this, further evaluation using a diverse and representative inference dataset, along with techniques like cross-validation, is recommended to gain a comprehensive understanding of the model’s generalization capabilities. If the 100% accuracy persists, considerations of practical implications and potential limitations in real-world scenarios are essential. This study only involved image interpretation of individual serovar at screening dilution with binary output. Further application of the trained model for leptospirosis diagnosis will require a more comprehensive dataset on panel serovars reactions with subsequent serum titration for quantitative result reporting.

## Conclusion

The developed model for leptospirosis detection showcases strong performance, reflected in precision, recall, and F1-Score metrics during training and testing. Precision at 0.81 indicates accurate identification of positive cases, while a recall score of 0.93 demonstrates the model’s effectiveness in capturing most leptospirosis instances. The balanced F1-Score of 0.87 emphasises the model’s efficiency in both precision and recall, crucial for reliable disease prediction. However, when tested with a separate dataset, 100% accuracy was achieved, raising concerns about potential over-fitting. To address this, adjustments to the model and thorough investigations into FP are recommended. Further considerations include adapting decision thresholds based on clinical needs, validating the model on diverse datasets, and assessing potential limitations in real-world scenarios. The comprehensive evaluation aims to optimise the model for reliable performance across various contexts.

## Data Availability

All data produced in the present study are available upon reasonable request to the authors

## Ethical consideration

The study reviewed and approved by the National Medical Research Registry (permit no: NMRR-16-1000-30876) according to law and research conduct in Malaysia. The archived image was obtained from routine diagnostic image acquisition from Zoonotic Laboratory, Bacteriology Unit, Institute for Medical Research, Malaysia.

## Acknowledgement

The authors would like to acknowledge Dr. Ami Fazlin Syed Mohamed, the director of Institute for Medical Research (IMR), Malaysia for the data support and image acquisition via the Bacteriology Unit. Special acknowledgement to Dr. Rohaidah Hashim, Head of Bacteriology Unit, and laboratory technicians of Zoonotic Laboratory, IMR, for their direct and indirect contribution to the study.

## Funding

The author(s) received no financial support for the research, authorship, and/or publication of this article. Data transfer agreement between the authors and IMR was acquired and granted.

## Conflicts of Interest

The authors declare no competing interests. The funders had no role in the writing of the manuscript, or in the content of the review

## Declaration of Generative AI and AI-assisted technologies in the writing process

During the preparation of this work the author(s) used Grammarly Inc. to improve the readability of the manuscript. After using this tool/service, the author(s) reviewed and edited the content as needed and take(s) full responsibility for the content of the publication.

## Notes

### Competing Interest Statement

The authors have declared no competing interest.

### Funding Statement

This study did not receive any funding

### Author Declarations

Ethics committee/IRB of the National Medical Research Registry (permit no: NMRR-16-1000-30876) gave ethical approval for this work

